# Estimating the disease burden of respiratory syncytial virus (RSV) in older adults in England during the 2023/24 season: a new national hospital-based surveillance system

**DOI:** 10.1101/2025.04.17.25325639

**Authors:** Rebecca Symes, Suzanne H. Keddie, Jemma Walker, Tricia McKeever, Shazaad Ahmad, David Arnold, Cariad M Evans, Emanuela Pelosi, Najib Rahman, Elizabeth Sapey, Maria Zambon, Conall Watson, Jamie Lopez Bernal, Wei Shen Lim, the HARISS network collaborators

## Abstract

**Background:** Respiratory syncytial virus (RSV) is an important cause of acute respiratory infection (ARI) in older adults. Vaccines that protect against severe RSV infection are now available.

**Aim:** We aimed to describe the incidence, presentation, severity and clinical outcomes of RSV-associated ARI in hospitalised older adults using a new Hospital-based ARI Sentinel Surveillance (HARISS) system in England in the winter prior to RSV vaccine introduction.

**Methods:** Adults aged ≥65 years from seven hospitals admitted for ≥24 hours with symptomatic ARI were included. Three groups were identified: RSV positive; influenza positive; negative for RSV, influenza and SARS-CoV-2. We estimated the hospitalisation rate of RSV-associated ARI compared to influenza-associated ARI and assessed clinical outcomes using Poisson regression and mortality using Cox regression across groups.

**Results:** This surveillance study included 2743 adults. During the 2023/4 season the hospitalisation rate for RSV-associated ARI was 58.3 per 100,000, compared to 114.6 per 100,000 for influenza-associated ARI. Hospitalisation rates increased with age. Exacerbation of chronic illness including lung disease, heart disease or frailty was a frequent cause of admission in RSV-associated ARI, with a combined incidence of 33.1 per 100,000. The majority of adults with RSV-associated ARI had at least one comorbidity (81%); a high proportion with immunosuppression (26%). Symptoms and clinical outcomes including mortality were similar between RSV- and influenza-associated ARI; 30-day mortality 10.6% vs 8.7% (adjusted hazard ratio 0.85,95% confidence interval 0.6-1.2).

**Conclusion:** In England, RSV infection is a common cause of hospitalisation in older adults. Symptoms at presentation, severity and clinical outcomes, including mortality, are comparable to influenza.

## Introduction

Respiratory syncytial virus (RSV) is a common seasonal respiratory virus that is well known for its high disease burden in infants. RSV is also an important cause of acute respiratory infection (ARI) in older adults, particularly in those living with frailty and comorbidities[1]. The global RSV-associated mortality rate in those aged over 70 years surpassed that in children under five years in 2019[2]. However, testing for RSV in older adults is not as widespread as testing for influenza or SARS-CoV-2 and the disease burden in adults may be underestimated using existing routine surveillance methods.

Previous UK modelling studies have estimated 156 hospitalisations per 100,000 persons (range 119-192) and 88 deaths per 100,000 persons (range 64-119) from RSV-attributable respiratory disease in adults ≥65 years during an average season[3] and an annual hospital admission rate of 71 and 251 per 100,000 persons in adults aged 65-74 years and 75 years and above respectively[4]. However, these estimates are from ecological studies and reflect data prior to the COVID-19 pandemic which is known to have impacted RSV epidemiology[5].

From 1 September 2024 an RSV immunisation programme for older adults was introduced in England[6]. The programme offers a single dose of bivalent RSV pre-F vaccine (Abrysvo, Pfizer) to all adults turning 75 years old alongside a one-off catch-up campaign for those aged 75 to 79 years old. To inform future RSV immunisation policy, it is pivotal to understand the burden and impact of RSV disease in older adults in England.

Therefore, a new enhanced Hospital-based Acute Respiratory Sentinel Surveillance (HARISS) system was introduced during the 2023/24 winter season with participating sites performing more consistent diagnostic testing for RSV, influenza and SARS-CoV-2 among older adults admitted to hospital that meet agreed case definitions for ARI. Surveillance data from HARISS has informed and will continue to inform the RSV vaccination programme in England.

This study aimed to describe the incidence, presentation, severity and clinical outcomes of RSV-associated ARI in hospitalised older adults. Specifically, this study estimates the incidence of RSV-associated hospitalisation in older adults compared to influenza-associated hospitalisation. Symptoms, severity of illness and clinical outcomes were compared in older adults hospitalised with ARI who tested positive on admission for RSV or influenza, or who tested negative for RSV, influenza and SARS-CoV-2.

## Methods

### Design and setting

This study involved a national, hospital-based, enhanced surveillance system (HARISS) across seven sentinel National Health Service (NHS) secondary care sites in England during winter 2023/4. Sites within the surveillance network include geographical representation across England (Supplementary Figure 1). One site contributed RSV case data only.

### Cohort

All HARISS sites were advised to test for RSV, influenza (A and B) and SARS-CoV-2 in patients aged ≥65 years admitted to hospital (for ≥24 hours) where the reason for admission was symptomatic ARI. Testing had to take place within 48 hours of admission and patients admitted for at least 24 hours to be included in the study. Symptomatic ARI included patients presenting with the following suspected diagnoses on admission:

1. Pneumonia, or pneumonitis
2. Non-pneumonia lower respiratory infection, or acute bronchitis
3. Exacerbation of chronic lung disease e.g. chronic obstructive pulmonary disease
4. Exacerbation of chronic heart disease e.g. heart failure
5. Exacerbation of frailty, or poor mobility e.g. fall
6. ARI with another reason for admission

For RSV cases, admissions included were between October 30, 2023 and February 11, 2024 (epidemiological week 44 to 6) and for influenza cases between November 27, 2023 and March 31, 2024 (epidemiological week 48 to 13) to capture seasonal cases during periods of peak activity. Test-negative cases (negative for RSV, influenza and SARS-CoV-2) were admitted between October 30, 2023 and March 31, 2024 (epidemiological week 44 to 13).

### Laboratory analysis

Testing pathways were site specific. Nasopharyngeal or combined nose and throat swabs were obtained as part of routine clinical management and tested by validated diagnostic platforms at each site. Testing consisted of molecular diagnostic assays including polymerase chain reaction (PCR) testing conducted either in a laboratory setting or at the point of care e.g. in the Emergency Department, by qualified personnel. Diagnostic platforms used were Cepheid GeneXpert, BioFire system, Hologic Panther Fusion, Roche cobas liat, Abbott ID NOW, Alinity M, Qiagen:QIAstat-Dx and in-house PCR respiratory panels.

### Data Collection

An electronic data capture form was completed for all known RSV and influenza cases and a sample of patients who tested negative for RSV, influenza and SARS-CoV-2. Negative cases were sampled in proportion to the number of RSV and influenza cases by week, across the surveillance period, i.e. larger numbers were sampled during weeks of higher RSV and influenza activity.

The data capture form collected enhanced surveillance information on cause of admission (multiple causes selected if applicable), symptoms on admission, antiviral use on admission, and in-hospital clinical outcomes using the Snap Survey platform[7] (Supplementary file).

Medical records were reviewed by trained staff employed at each site. Clinical outcomes were captured at 30 days after sample date, or at discharge, whichever was sooner.

### Linked data sets

The Immunisation Information System (IIS) is a national vaccine register containing demographic information on the population of England registered with a General Practitioner. IIS was accessed for sex, ethnicity, index of multiple deprivation (IMD), influenza/COVID-19 risk group status (comorbidities), and death date. Comorbidities were identified from the NHS CaaS (Cohorting as a Service)[8], developed for call-recall purposes to identify at risk individuals requiring influenza or COVID-19 vaccine based on primary care electronic health records. Hospital Episode Statistics (HES) is the national electronic database of hospital admissions, outpatient appointments and emergency department attendances at NHS hospitals in England[9]. HES was accessed to calculate length of stay.

### Analysis

Demographic, coinfection, comorbidities, symptoms, and clinical outcomes were described for RSV, influenza and negative cases. Mann-Whitney U tests and Pearson’s chi-squared tests were used to compare demographic and comorbidities characteristics between groups.

Hospitalisation rates were estimated using NHS trust catchment populations produced by the Office for Health Improvement and Disparities. Catchment populations from 2019 were used as the latest figures not subject to COVID-19 pandemic changes in hospital activity[10].

Poisson regression was used to estimate the relative risk of each clinical outcome in RSV-associated ARI compared to influenza-associated ARI and test-negative ARI. To compare mortality between groups hazard ratios were estimated using Cox regression. Death outcomes considered were death during admission due to ARI at 30 days or discharge (whichever was sooner), and all-cause mortality at 30, 60 and 90 days. Models were adjusted for age, sex, ethnicity, IMD, geographic region and presence of at least one comorbidity. Antiviral use prior to admission, and influenza vaccination status were not included in the final models as they made no difference to the relative risks or hazard ratios. We regarded p-values <0.05 statistically significant.

For cases who were admitted with the same pathogen within 30 days, only the first admission was included. For cases admitted with separate RSV and influenza episodes, these were treated as independent episodes if the admission dates were at least 14 days apart. Cases were excluded from the clinical outcome and mortality analyses if they had coinfection or if demographic characteristics included in the model were not available.

Analyses were completed using R Studio 4.3.2 and Stata 18[11,12].

## Results

### 1. Cases

Between 30 October 2023 and 11 February 2024, 720 adults aged ≥65 years were reported as testing RSV positive within 48 hours of admission across seven hospital sites. Peak RSV hospitalisations occurred during December 2023 (Figure 1); ARI was the primary reason for admission or contributed to admission in 611 cases (84.9%) (Figure 2).

**Figure 1:**
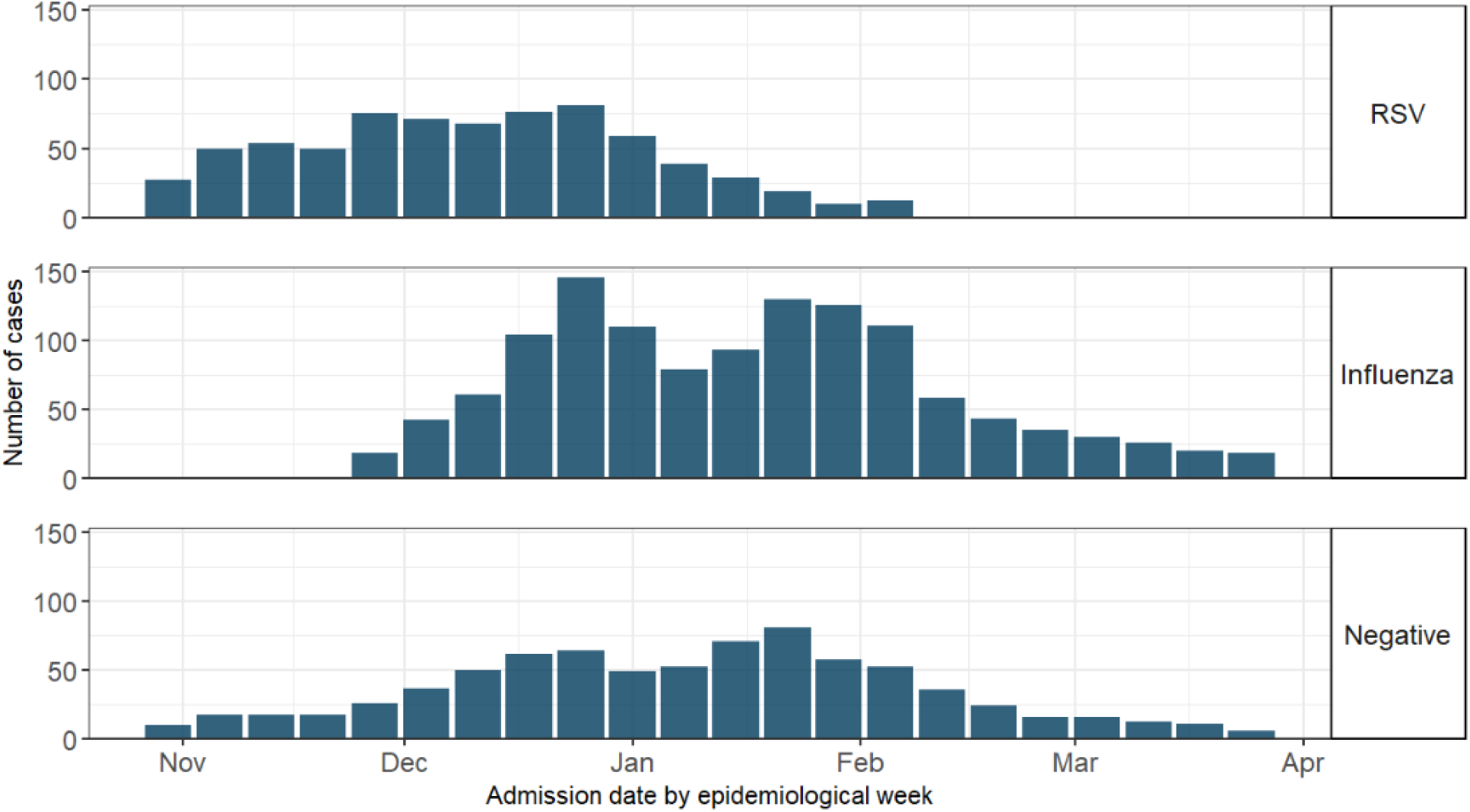
Epidemiological curve of all included cases.

**Figure 2:**
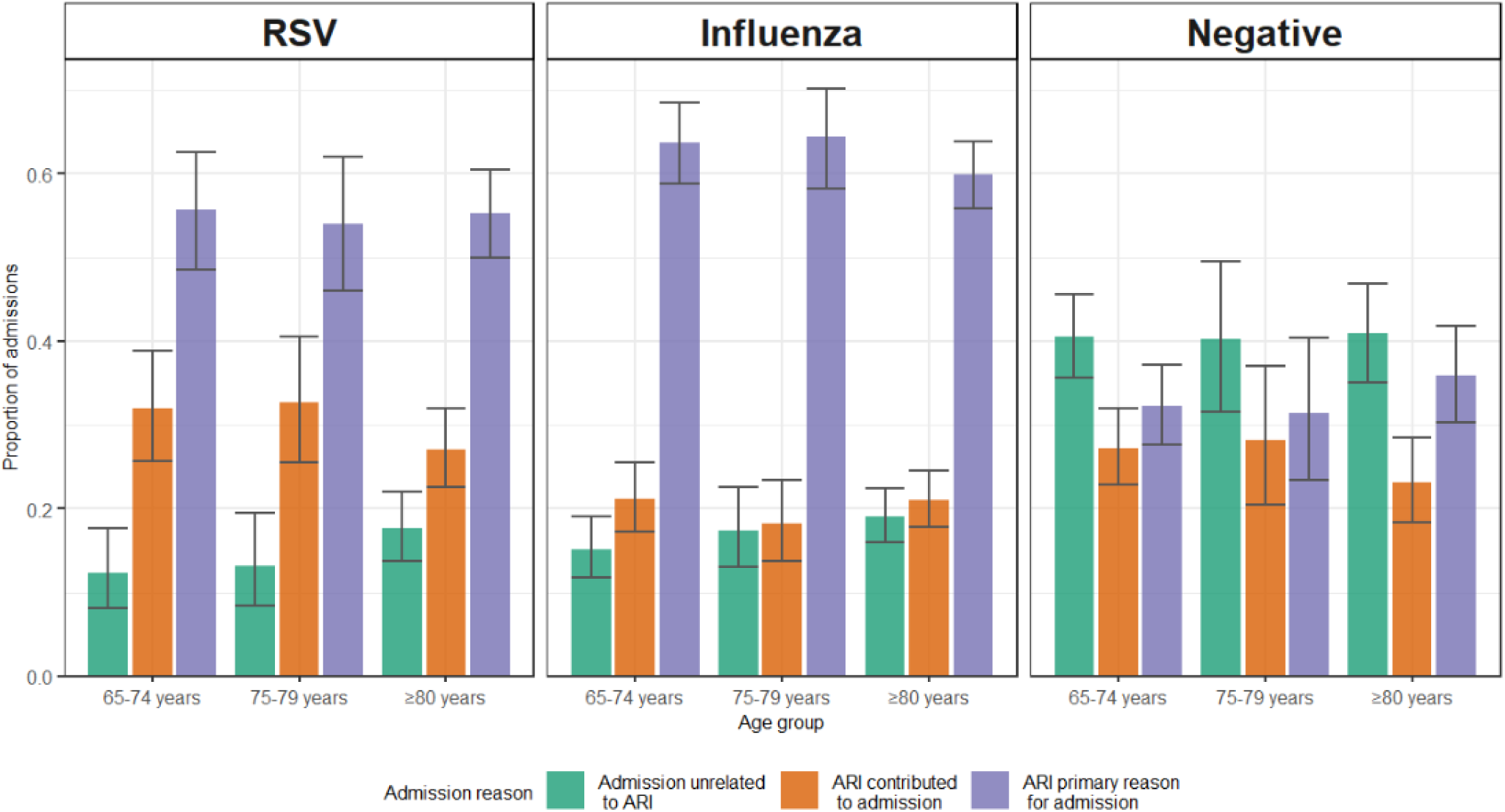
Admission reason for RSV, influenza and negative cases across age groups, with 95% confidence intervals.

Between 27 November 2023 and 31 March 2024, 1250 adults aged ≥65 years were reported as testing influenza positive (A or B) within 48 hours of admission across six sites. Peak influenza-positive hospitalisations occurred in the last week of December 2023 and again during the end of January 2024 (Figure 1); ARI was the primary reason for admission or contributed to admission in 1032 cases (82.6%) (Figure 2).

Of the 790 hospitalised cases sampled that tested negative for RSV, influenza and SARS-CoV-2 within 48 hours of admission, ARI was the primary reason for admission or contributed to admission in 469 cases (59.4%) (Figure 2).

### 2. Characteristics of ARI cases

Of cases where ARI was the primary reason for admission or contributed to admission (n=2112), the median age in RSV-associated ARI was 80 years (IQR 73-86 years). The majority (84%) identified as White British ethnicity and 30% were in the most deprived IMD quintile. A high proportion had at least one comorbidity (81%); 57% chronic heart or vascular disease, 47% chronic lung disease, 26% immunosuppressed (Table 1). In the 65-74- and 75-79-year age groups a higher proportion of cases had at least one comorbidity (82% and 84% respectively), compared to 78% in the ≥80-year age group. Chronic respiratory disease and immunosuppression were more common in younger age groups, compared to the ≥80-year age group (56% and 57% vs 37% respectively for chronic respiratory disease and 34% and 29% vs 19%, respectively for immunosuppression) (Supplementary Table 1).

**Table 1:** Characteristics of RSV-associated ARI, influenza-associated ARI and test-negative ARI cases.

|  | RSV-associated<br>ARI<br>n=611 |  | Influenza-associated<br>ARI<br>n=1032 |  | p-value*<br>RSV vs<br>influenza | Test-negative<br>ARI<br>n=469 |  | p-value**<br>RSV vs<br>negative |
| --- | --- | --- | --- | --- | --- | --- | --- | --- |
| **Age**<br>**Median (IQR)** | 80 | (73-86) | 79 | (73-85) | 0.185 | 75 | (71-84) | <0.001 |
| **Sex, female (%)** | 363 | (59) | 560 | (54) | 0.047 | 239 | (51) | 0.006 |
| **Ethnicity, n (%)** |  |  |  |  | 0.001 |  |  | 0.026 |
| White - British | 515 | (84) | 799 | (77) |  | 372 | (79) |  |
| White - Other | 32 | (5) | 69 | (7) |  | 39 | (8) |  |
| Mixed or multiple ethnic groups | 0 | (0) | 7 | (1) |  | 4 | (1) |  |
| Asian or Asian British | 28 | (5) | 86 | (8) |  | 21 | (4) |  |
| Black, Black British, Caribbean or African | 7 | (1) | 27 | (3) |  | 11 | (2) |  |
| Other ethnic group | 10 | (2) | 15 | (1) |  | 5 | (1) |  |
| **Deprivation (IMD quintile), n (%)** |  |  |  |  | 0.002 |  |  | 0.422 |
| 1 (Most deprived) | 181 | (30) | 371 | (36) |  | 133 | (28) |  |
| 2 | 95 | (16) | 186 | (18) |  | 89 | (19) |  |
| 3 | 101 | (17) | 170 | (16) |  | 79 | (17) |  |
| 4 | 93 | (15) | 148 | (14) |  | 78 | (17) |  |
| 5 (Least deprived) | 136 | (22) | 155 | (15) |  | 88 | (19) |  |
| **Comorbidities, n (%)** |  |  |  |  |  |  |  |  |
| At least one comorbidity | 492 | (81) | 900 | (87) | <0.001 | 376 | (80) | 0.946 |
| Chronic respiratory disease | 287 | (47) | 509 | (49) | 0.384 | 228 | (49) | 0.636 |
| Chronic heart disease & vascular disease | 350 | (57) | 602 | (58) | 0.715 | 264 | (56) | 0.791 |
| Chronic kidney disease | 168 | (27) | 314 | (30) | 0.228 | 120 | (26) | 0.526 |
| Diabetes & other endocrine disorders | 164 | (27) | 334 | (32) | 0.021 | 137 | (29) | 0.428 |
| Chronic neurological disease | 159 | (26) | 364 | (35) | <0.001 | 137 | (29) | 0.273 |
| Immunosuppression | 157 | (26) | 208 | (20) | 0.011 | 110 | (23) | 0.438 |
| Morbid obesity | 31 | (5) | 42 | (4) | 0.406 | 28 | (6) | 0.612 |
| Chronic liver disease | 22 | (4) | 41 | (4) | 0.805 | 15 | (3) | 0.848 |
| Severe mental illness | 20 | (3) | 47 | (5) | 0.254 | 17 | (4) | 0.884 |
| Asplenia | 7 | (1) | 17 | (2) | 0.544 | 5 | (1) | 0.902 |
| **Number of comorbidity risk flags,**<br>**median (IQR)** | 2 | (1-3) | 2 | (1-3) | 0.034 | 2 | (1-3) | 0.817 |

\*Comparison of RSV-associate ARI and influenza-associated ARI cases using Chi-squared test for categorical variables and Mann-Whitney U test for continuous variables.
\*\*Comparison of RSV-associated ARI and test-negative ARI using Chi-squared test for categorical variables and Mann-Whitney U test for continuous variables.

In influenza-associated ARI, a lower proportion identified as White British ethnicity (77%), with 36% in the most deprived quintile. A higher proportion had at least one comorbidity (87%), however there were similar proportions of chronic respiratory and chronic heart and vascular disease between groups. There were lower levels of immunosuppression (20%) compared to RSV-associated ARI. The test-negative ARI group was younger (median age 75). There were similar levels of comorbidities compared to RSV-associated ARI.

Coinfection with another respiratory pathogen was reported in 5% (n=32) of RSV-associated ARI cases and in 5% (n=56) of influenza-associated ARI cases.

### 3. Incidence of RSV- and influenza-associated ARI hospitalisation

Of the 611 older adults hospitalised with RSV-associated ARI, 178 (29.1%) were aged 65-74 years, 138 (22.6%) 75-79 years and 295 (48.3%) ≥80 years; corresponding figures for older adults hospitalised with influenza-associated ARI (n=1032) were 337 (32.7%), 218 (21.1%) and 477 (46.2%).

Between 30 October 2023 and 11 February 2024, the incidence of hospitalisation from RSV-associated ARI was 58.3 per 100,000 in adults ≥65 years. Incidence showed an increasing trend from the youngest (65-74 years, 30.9 per 100,000) to oldest age group (≥80 years, 107.9 per 100,000). In comparison, the incidence of hospitalisation from influenza-associated ARI was 114.6 per 100,000 in adults ≥65 years between November 27, 2023 and March 31, 2024. Similarly, incidence showed an increasing trend from the youngest (65-74 years, 68.1 per 100,000) to oldest age group (≥80 years, 203.3 per 100,000). There was variation in the incidence of both RSV-associated ARI and influenza-associated ARI between hospital sites (Table 2).

**Table 2:**
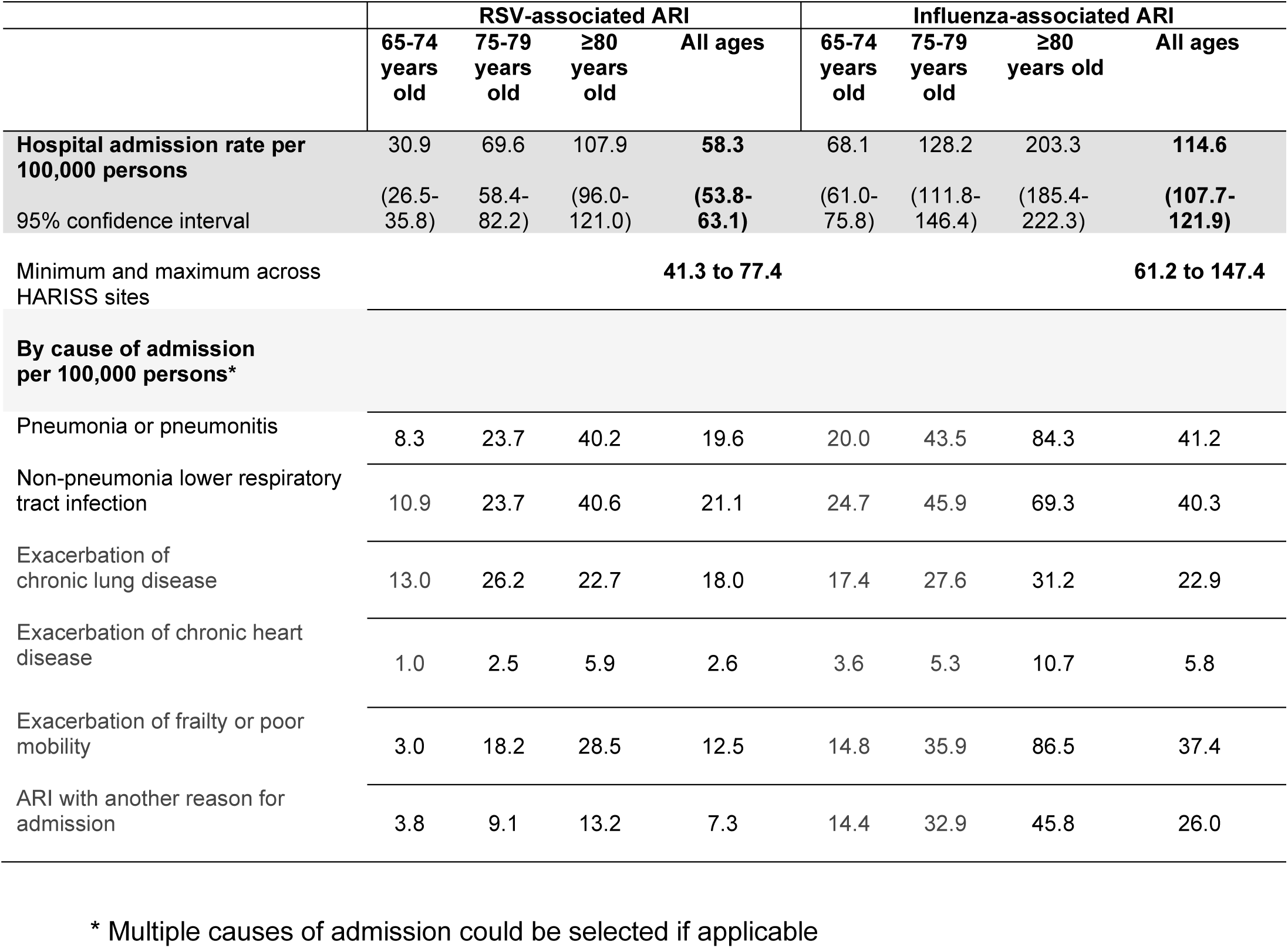
Incidence of RSV- and influenza-associated ARI hospitalisation.

In cases with RSV-associated ARI, exacerbation of chronic lung disease was the most common cause of admission in the 65-74 years and 75-79 years age groups; while non-pneumonia lower respiratory infection was the most frequent cause of admission in cases ≥80 years. In influenza-associated ARI cases, non-pneumonia lower respiratory infection was the commonest cause of admission in patients aged 65-74 and 75-79 years. Exacerbation of frailty or poor mobility had the highest incidence in those aged ≥80 years (Table 2).

### 4. Symptoms and signs on admission in ARI cases

The three commonest symptoms and signs in RSV-associated ARI were new or increased cough (77%), shortness of breath (76%) and new or increased sputum volume (43%). The three commonest symptoms in influenza-associated ARI were new or increased cough, shortness of breath and fever. The relative frequency of these symptoms and signs was similar for test-negative ARI cases (Figure 3).

**Figure 3:**
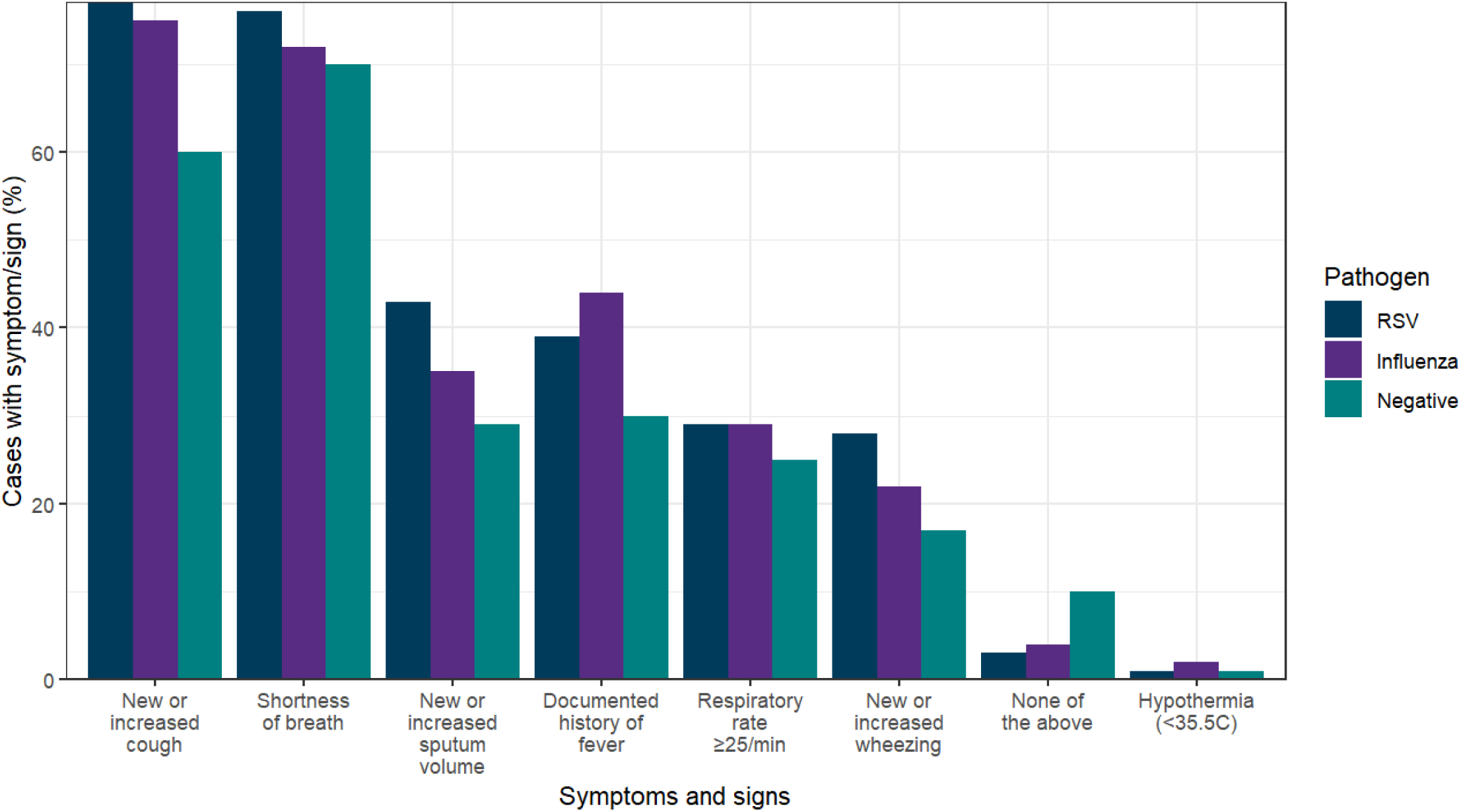
Symptoms and signs in RSV-associated ARI, influenza-associated ARI and test-negative ARI cases.

### 5. Clinical outcomes and mortality in ARI cases

Models were adjusted for age, sex, ethnicity, IMD, region and presence of at least one comorbidity. In RSV-associated ARI, 68.8% required oxygen therapy. The relative risk (RR) was not significantly different after adjustment when compared to influenza-associated ARI or test-negative ARI. Although the proportion of patients receiving high flow nasal oxygen, non-invasive ventilation or continuous positive airway pressure was higher in RSV-associated ARI than influenza-associated ARI (7.1% vs 5.5%) there was not a significant difference in relative risk in the adjusted model (adjusted RR 1.21 (0.79,1.84)). Only seven RSV-associated ARI cases (1.2%) were admitted to intensive care (ITU). There was no significant difference in relative risk of ITU admission after adjustment in RSV-associated ARI vs influenza-associated ARI (adjusted RR 0.45 (95% CI 0.20,1.06) but there was significantly higher ITU admissions in test-negative ARI when compared to RSV-associated ARI (adjusted RR 0.33 (95% CI 0.13,0.85) (Table 3).

**Table 3:**
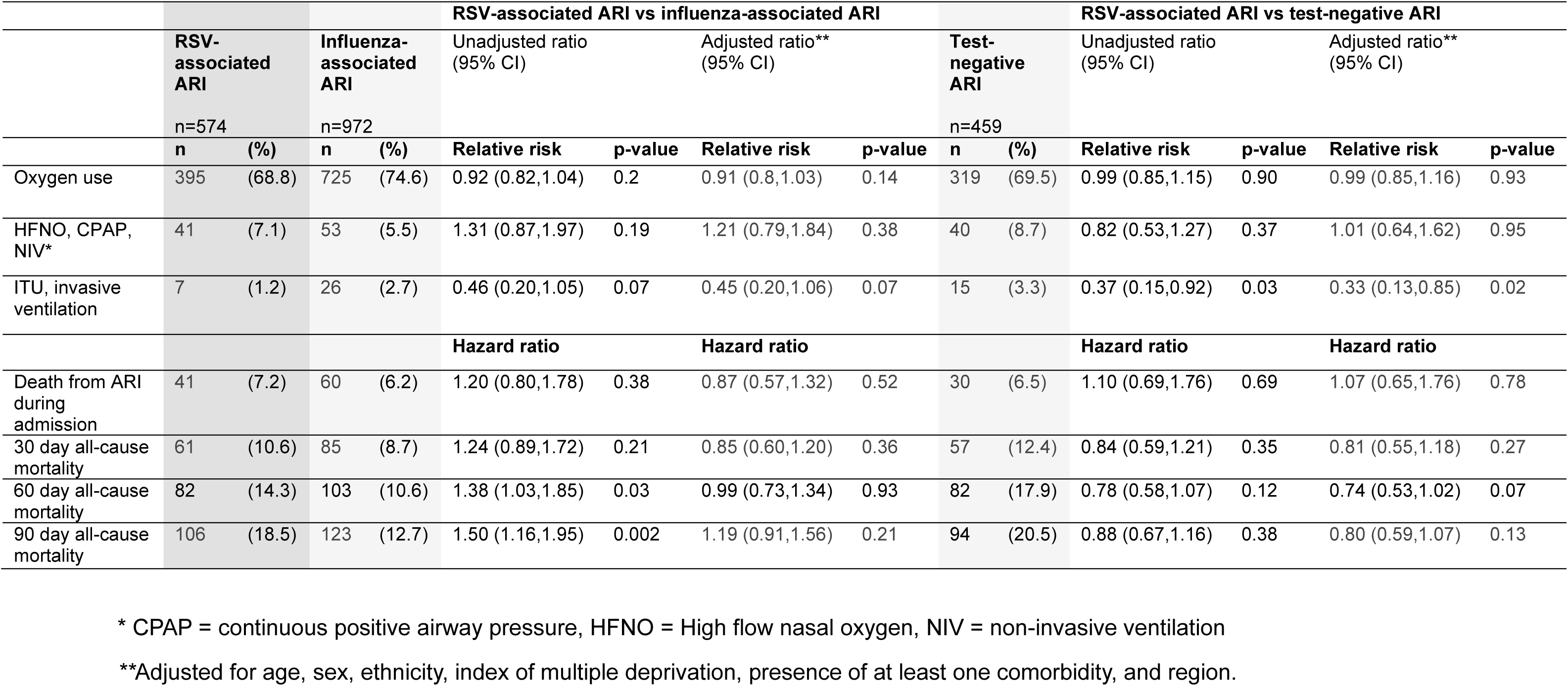
Comparison of clinical outcomes and mortality in RSV-associated ARI cases vs influenza-associated ARI and test-negative ARI cases.

There were no differences detected in in-hospital mortality from ARI and all-cause mortality at 30 and 60 days in RSV-associated ARI compared to influenza-associated ARI or test-negative ARI. At 90 days the unadjusted hazard ratio (HR) showed that all-cause mortality was higher in RSV-associated ARI compared to the influenza-associated ARI (unadjusted HR 1.50 (95% CI 1.16,1.95)) but after adjustment there was no difference (adjusted HR 1.19 (95% CI 0.91,1.56) (Table 3). All-cause mortality at 30, 60 and 90 days in RSV-associated ARI was higher in persons aged ≥80-years old compared to those aged 65-74 and 75-79 years (30-day all-cause mortality was 6.7%, 7.6% and 14.4% in persons aged 65-74, 75-79 and ≥80 years respectively) (Supplementary Table 2).

RSV-associated ARI cases had a median length of stay of 6 days (IQR 3-11). Length of stay was the same in influenza-associated ARI cases and test-negative ARI cases, both with a median of 6 days (IQR 3-11).

## Discussion

### Main findings

Data from the newly established HARISS surveillance system indicate a substantial hospital burden of RSV-associated ARI in older adults in England. The incidence of RSV-associated ARI hospitalisation in a well-tested older adult population was around half of that of influenza-associated ARI. Much of the RSV burden was due to exacerbation of existing chronic illness, including chronic respiratory disease, chronic heart disease and frailty. There are high levels of comorbidities in those admitted to hospital with ARI, with higher levels of immunosuppression in those admitted with RSV in comparison to influenza. Symptoms and signs on presentation were difficult to differentiate between pathogen groups. Clinical severity outcomes and mortality were similar between RSV- and influenza-associated ARI. RSV-associated ARI hospitalisation and mortality increased with increasing age.

### Comparison to existing literature

In line with existing literature from England, RSV is demonstrated in this surveillance study to account for considerable seasonal ARI hospital admissions in older adults[3.4]. Previous ecological studies reflect epidemiology several years ago, prior to the COVID-19 pandemic. This surveillance study from winter 2023/4 corroborates previous evidence on disease burden and provides current estimates of hospitalisation rates in the season prior to RSV vaccine rollout, enabling the monitoring of RSV activity as vaccination is introduced.

In comparison to previous modelling studies in England[3,4] our estimated hospital admission rates from this surveillance study are lower (Fleming *et al.* report RSV-attributable hospitalisations among adults ≥65 years in the UK of 156/100,000 between September and mid-May)[3]. However, there are key differences in methodology to be considered. We included only patients who were tested within 48 hours of admission, were admitted for ≥24 hours, and were reported as being admitted due to symptomatic ARI. Our surveillance period of interest focused on periods of highest RSV and influenza activity to obtain detailed clinical information, as opposed to an entire winter season or calendar year. Additionally, changes in hospital admission thresholds over time have been reported[13].

Our RSV-associated ARI hospital admission rate in adults ≥65 years of 58.3 per 100,000 is higher than the UKHSA SARI-Watch surveillance system[14] hospital admission rate for confirmed RSV cases during the same time period (51.5 per 100,000). These differences highlight the importance of more consistent testing and likely under-reporting due to limited testing in some hospital settings.

Compared to other European studies our hospital admission rates are considerably higher than those reported by Martinón-Torres *et al.* in Spanish adults ≥65 years (23.7 per 100,000), where more limited diagnostic testing is discussed[15]. Our estimates are more in line with those recently reported by Vera-Punzano *et al.* where routine testing for RSV and influenza was used. Although the four-year average hospitalisation rate is higher at 84.7 per 100,000 our incidence is very similar to the 59.7 per 100,000 reported hospitalisation rate during one season[16].

We observed variation in the reported hospital admission rate in adults aged ≥65 years across HARISS sites, ranging from 41 to 77 per 100,000 in RSV-associated ARI, and 61 to 147 per 100,000 in influenza-associated ARI. This may reflect differences in admission thresholds, testing practices and case mix. Regional variation of respiratory infection in England has been reported. For example, higher rates of GP consultations for lower respiratory tract have been reported in the North and Midlands regions of England and in urban and deprived areas[17].

We report high levels of comorbidities in those admitted with RSV-associated ARI, especially in those aged 65-79 years. Chronic cardiac and vascular disease was the most common comorbidity in our cohort consistent with reports that cardiovascular disease is associated with hospitalisation in 45% to 63% of adults with confirmed RSV[18]. The comparatively high proportion of immunosuppression in the RSV group is in line with other studies comparing RSV to influenza[19]. A significant burden of RSV-related disease in adults with immunosuppression is reported[20,21]. This study shows particularly high levels of at least one comorbidity, chronic respiratory disease and immunosuppression in adults aged 65-74 and 75-79 years which is aligned with evidence suggesting the presence of chronic conditions as well as immunosenescence due to ageing are factors that contribute to severe RSV outcomes[22].

A high rate of hospitalisation was caused by exacerbation of existing chronic illness, which may be under-recognised in hospital settings where testing is more restricted. RSV is a major cause of COPD and asthma exacerbations[23] and admission for congestive heart failure has been attributed to RSV[24]. This study also illuminates a significant burden of disease in older adults presenting with exacerbation of frailty and poor mobility, particularly in those ≥80 years.

Previous studies examining the symptom profile of adults hospitalised with RSV or influenza infection have reported more fever in those presenting with influenza, and more symptoms/signs of cough, dyspnoea and wheeze in adults with RSV[25,26]. In this study, the proportion of patients experiencing each symptom did not differ significantly between groups. This could reflect differences in study cohort age, with our study reporting a higher median age than the studies described. Concordant with our findings, a large cohort study focusing on older adults reported similar symptoms and signs in patients admitted with RSV or influenza[24]. The poor discriminatory value of symptoms in older adults to distinguish between pathogens highlights the importance of consistent testing for respiratory pathogens.

After adjustment, no significant differences were observed in clinical outcomes or mortality between patients admitted with RSV- and influenza-associated ARI. This is consistent with other European studies reporting clinical outcomes of RSV and influenza to be similar once hospitalisation has occurred[27,28,29] and similar 30-day mortality between these pathogens[24,27,30]. We recognise clinical outcomes of influenza may be influenced by antiviral treatment or prior vaccination. Our observed mortality in RSV-associated ARI cases of 7.2% during admission is comparable to the 7.4% mortality reported by Clausen *et al.*[29] and our 30-day mortality of 10.6% is comparable to the 10% mortality reported by Boon *et al.*[27].

In addition, this study showed considerable severity and mortality in ARI patients testing negative for RSV, influenza, and SARS-CoV-2. There is an increasing recognition of the significant morbidity and mortality from other respiratory pathogens such as hMPV and rhinovirus in older adults and it is reasonable to suggest these have contributed to the burden observed in the negative group[4,27].

This surveillance study expands current knowledge by providing up-to-date hospitalisation rates for RSV-associated ARI in older adults in England. Consistent testing of older adults admitted with a broad range of ARI presentations has provided insight into an under-recognised burden of RSV disease.

### Strengths and limitations

This study includes data from a single winter season. The season showed moderate levels of influenza activity in England[31]. There is the potential for under-ascertainment due to use of nasopharyngeal/combined nose and throat swabbing only and imperfect sensitivity of testing approaches recognised in respiratory virus testing[32]. Sites were recruited to HARISS due to consistent testing policies for RSV, influenza and SARS-CoV-2 in patients presenting with symptomatic ARI. However, as testing was performed for routine clinical care and not research purposes there may be some variation in testing procedures due to differences in local protocols and staff practices.

There are notable strengths to this study including the use of a national, muti-center, surveillance system in England with wide geographical representation. Sites in the HARISS network aimed to test all older adults presenting with ARI, including those who presented with exacerbations of chronic conditions. Patient notes were directly reviewed to provide accurate information. The direct review of patient notes, alongside the stringent inclusion criteria of respiratory testing within 48 hours of admission, resulted in the inclusion of only patients who had been admitted due ARI, as opposed to an incidental finding after admission for another cause.

### Conclusion and implications

RSV-associated ARI is an important cause of hospitalisation and mortality in adults ≥65 years in England, with the highest rates of hospitalisation and mortality in those aged ≥80 years. The lack of a characteristic symptom profile for RSV-associated ARI underscores the importance of consistent testing pathways for adults hospitalised with ARI to detect RSV infection in addition to influenza and SARS-CoV-2. Given the frequency and severity of illness associated with RSV infection in older adults, the potential for public health benefits from RSV immunisation programmes are likely to be substantial.

## Data Availability

Original data are confidential and no additional data available.

## HARISS network collaborators

Elisabeth North^a^, Matthew Donati^b^, Simon Tazzyman^c^, Thushan de Silva^d^, Tristan Clark^e^, Nicola White^e^, Tine Panduro^f^, Monique Andersson^g^, Suzy Gallier^h,I,j,k^, Sowsan Atabani^l^, Timothy Felton^m,n^, Louise Berry°, Louise Lansbury^p,q^, Christopher Rawlinson^r^

a. Academic Respiratory Unit, North Bristol NHS Trust, Bristol, UK
b. UK Health Security Agency, South West Regional Laboratory and Severn Infection Sciences, Bristol, UK
c. South Yorkshire and Bassetlaw Pathology Network, Sheffield Teaching Hospitals NHS Trust, Sheffield, UK
d. Division of Clinical Medicine, School of Medicine and Population Health, The University of Sheffield, Sheffield, UK
e. University Hospital Southampton, NHS Foundation Trust, Southampton, UK
f. Oxford Respiratory Trials Unit, University of Oxford, Oxford, UK
g. Nuffield Department of Clinical Laboratory Sciences, Radcliffe Department of Medicine, University of Oxford, Oxford
h. National Institute for Health Research Birmingham Biomedical Research Centre, University of Birmingham, UK.
i. National Institute for Health Research Midlands Patient Safety Research Collaboration, University Hospital Birmingham NHS Foundation Trust, UK.
j. PIONEER Hub in acute care, Health Data Research UK
k. Institute of Inflammation and Ageing, University of Birmingham, Birmingham, UK
l. UKHSA Birmingham Public Health Laboratory, Heartlands Hospital, University Hospitals Birmingham NHS Foundation Trust, Birmingham, UK
m. Division of Infection, Immunity to Infection & Respiratory Medicine, The University of Manchester, Manchester, UK
n. Wythenshawe Hospital, Manchester University NHS Foundation Trust, Manchester, UK
o. Department of Microbiology, Nottingham University Hospitals NHS Trust, Nottingham, UK
p. Faculty of Medicine and Health Sciences, University of Nottingham, Nottingham, UK
q. National Institute for Health Research (NIHR) Nottingham Biomedical Research Centre, UK
r. UK Health Security Agency, London, UK

## Acknowledgements

We would like to acknowledge the following colleagues for their contributions to HARISS. *UKHSA Respiratory Virus Unit, London, UK*: Beatrix Kele, Tiina Talts, Katja Hoschler, Busayo Elegunde, Thomas Williams, Praveen Sebastianpillai. *UKHSA, London, UK*: Suzanne Elgohari. *Clinical Trails Assistant Team, Sheffield Teaching Hospitals NHS Trust, Sheffield, UK*: Hannah Dunn, Jessica Gregory, Christopher Norman, Cameron Obie, Abigail Olojede, Jennifer Rufus, Franciszek Skalbania, Fred Smith, Nicole Whitfield, Chloe Mathewman, Hannah Maughan, Georgie MacLoughlin, Nicola Tinker.

Department of Virology, Sheffield Teaching Hospitals NHS Trust, Sheffield, UK: Alex Yates *Oxford Respiratory Trials Unit, University of Oxford, Oxford, UK:* Melissa Dobson, Kathryn Lafferty, Jasmin Sadler-Ladell.

*University Hospital Southampton, NHS Foundation Trust, Southampton, UK:* Sophie Willis, Chibuzor Amechi.

*PIONEER Hub in acute care, Health Data Research UK*: Mohammed Ahmed, Alan Kwok, Felicity Evison.

*UKHSA Birmingham Public Health Laboratory, Heartlands Hospital, University Hospitals Birmingham NHS Foundation Trust, UK:* Judith Workman, Amanda Symonds.

*Manchester University NHS Foundation Trust, Manchester, UK*: Simona Serban, Maria Swizel Furtado e Aguiar, Manju Juby, Georgios Chalikias.

*Nottingham University Hospitals NHS Trust, Nottingham, UK:* Lesley Bendall, Depu Parakkalpapu, Michelle Lister, Benjamin Sloan.

*UK Health Security Agency, South West Regional Laboratory and Severn Infection Sciences, Bristol, UK:* Paul North, Rich Hopes.

We would like to acknowledge the NIHR Respiratory Biomedical Research Centres (Southampton, Nottingham, Manchester, Bristol and Oxford, and Sheffield), NIHR Respiratory Translational Research Collaboration at these sites, PIONEER Hub in acute care, Birmingham (NIHR Midlands Patient Safety Research Collaboration (PSRC) and NIHR Birmingham Biomedical Research Centre funded).

