## Supplementary file for "Estimating the disease burden of respiratory syncytial virus (RSV) in older adults in England during the 2023/24 season: a new national hospital-based surveillance system"

**Supplementary Figures & Tables**

**Supplementary Figure 1: Map of England with location of HARISS sites**

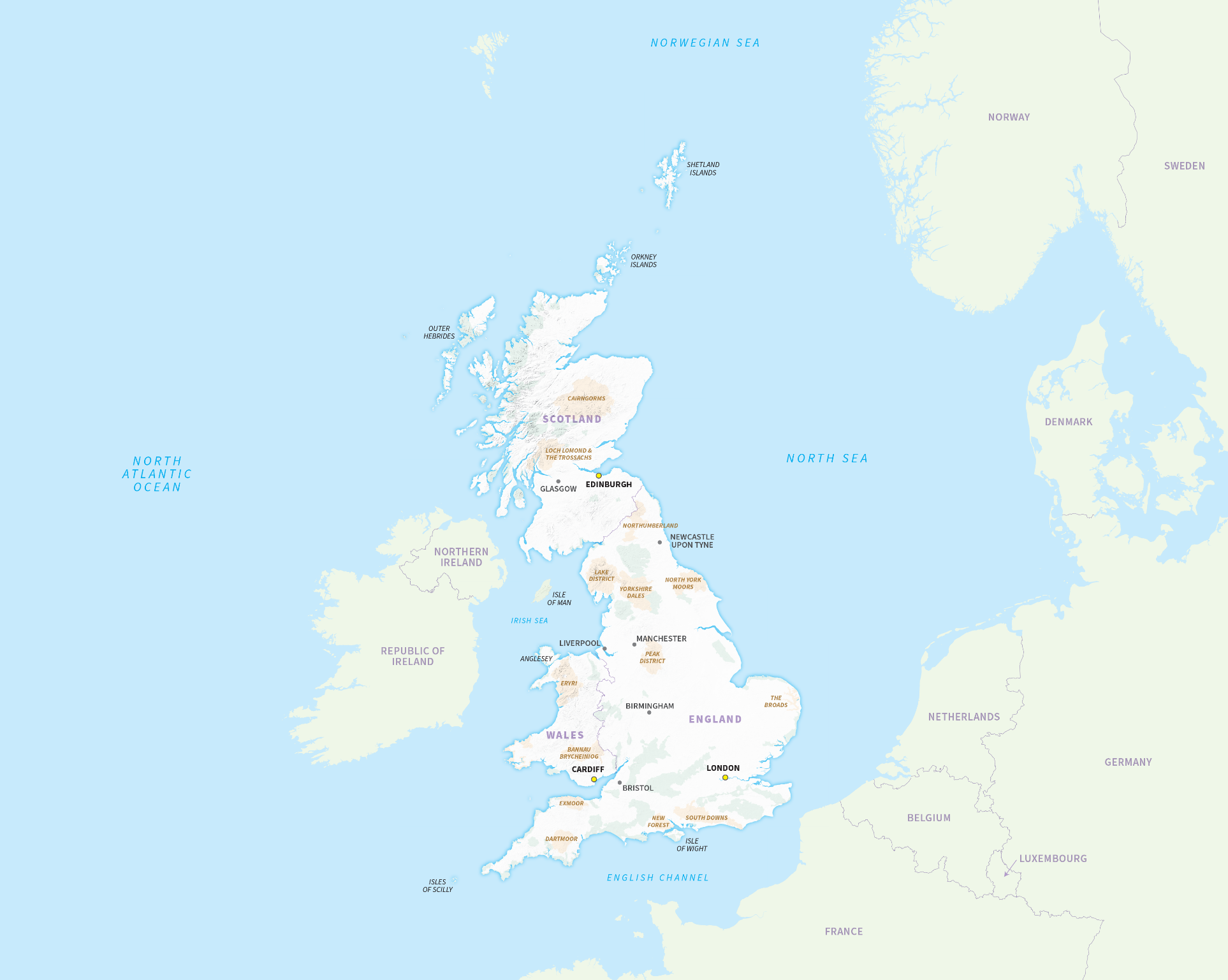

Acknowledgement: Ordnance Survey January 2025. Adapted to show HARISS sites location.

**Supplementary Table 1: Comorbidities in RSV-associated ARI by age group**

| n (%) | 65-74 years old  n=178 | | | | 75-79 years old  n=138 | | ≥80 years old  n=295 | |
| --- | --- | --- | --- | --- | --- | --- | --- | --- |
| At least one comorbidity | 146 |  | (82) |  | 116 | (84) | 230 | (78) |
| Chronic respiratory disease | 99 |  | (56) |  | 78 | (57) | 110 | (37) |
| Chronic heart disease and vascular disease | 89 |  | (50) |  | 83 | (60) | 178 | (60) |
| Chronic kidney disease | 26 |  | (15) |  | 36 | (26) | 106 | (36) |
| Diabetes and other endocrine disorders | 48 |  | (27) |  | 43 | (31) | 73 | (25) |
| Chronic neurological disease | 30 |  | (17) |  | 29 | (21) | 100 | (34) |
| Immunosuppression | 61 |  | (34) |  | 40 | (29) | 56 | (19) |
| Morbid obesity | 12 |  | (7) |  | 11 | (8) | 8 | (3) |
| Chronic liver disease | 9 |  | (5) |  | 7 | (5) | 6 | (2) |
| Severe mental illness | 5 |  | (3) |  | 7 | (5) | 8 | (3) |
| Asplenia | 7 |  | (4) |  | 0 | (0) | 0 | (0) |

**Supplementary Table 2:** **Severity outcomes and mortality in RSV-associated ARI by age group**

| n (%) | 65-74 years old  n=165 | | 75-79 years old  n=132 | | ≥80 years old  n=277 | |
| --- | --- | --- | --- | --- | --- | --- |
| Oxygen use | 117 | (70.9) | 89 | (67.4) | 189 | (68.2) |
| High-flow nasal oxygen | 2 | (1.2) | 6 | (4.5) | 12 | (4.3) |
| CPAP or NIV* | 8 | (4.8) | 10 | (7.6) | 10 | (3.6) |
| Invasive ventilation | 1 | (0.6) | 1 | (0.8) | 0 | (0) |
| Intensive care admission | 3 | (1.8) | 2 | (1.5) | 1 | (0.4) |
| ECMO** | 0 | (0) | 0 | (0) | 0 | (0) |
| Death from ARI during admission | 9 | (5.5) | 8 | (6.1) | 24 | (8.7) |
| 30 day all-cause mortality | 11 | (6.7) | 10 | (7.6) | 40 | (14.4) |
| 60 day all-cause mortality | 19 | (11.5) | 13 | (9.8) | 50 | (18.1) |
| 90 day all-cause mortality | 21 | (12.7) | 21 | (15.9) | 64 | (23.1) |

***** Continuous positive airway pressure or non-invasive ventilation

** Extracorporeal membrane oxygenation

**Supplementary Table 3: Odds of admission by cause in RSV-associated ARI vs influenza-associated ARI cases**

|  |  | **RSV**  **n(%)** | **Influenza**  **n(%)** | **Unadjusted odds ratio** | **p value** | **Adjusted**  **OR*** | **p value** |
| --- | --- | --- | --- | --- | --- | --- | --- |
| **Pneumonia or pneumonitis** | | 193 (33.6) | 348 (35.8) | 0.91 (0.73,1.13) | 0.39 | 0.89 (0.72,1.1) | 0.31 |
| **Non-pneumonia LRTI** | | 208 (36.2) | 340 (35.0) | 1.06 (0.85,1.31) | 0.62 | 1.05 (0.85,1.31) | 0.62 |
| **Exacerbation of**  **chronic lung disease** | | 178 (31.0) | 195 (20.1) | 1.79 (1.41,2.27) | <0.001 | 1.85 (1.46,2.36) | <0.001 |
| **Exacerbation of chronic heart disease** | | 26 (4.5) | 50 (5.1) | 0.87 (0.54,1.42) | 0.59 | 0.87  (0.54,1.42) | 0.59 |
| **Exacerbation of frailty or poor mobility** | | 120 (20.9) | 308 (31.7) | 0.59 (0.47,0.73) | <0.001 | 0.54 (0.42,0.70) | <0.001 |
| **Another reason for admission** | | 73 (12.7) | 226 (23.3) | 0.48 (0.36,0.64) | <0.001 | 0.48 (0.36,0.64) | <0.001 |

*Adjusted for age

**Supplementary Figure 2**

a) **Symptom upset plot for RSV-associated ARI cases**

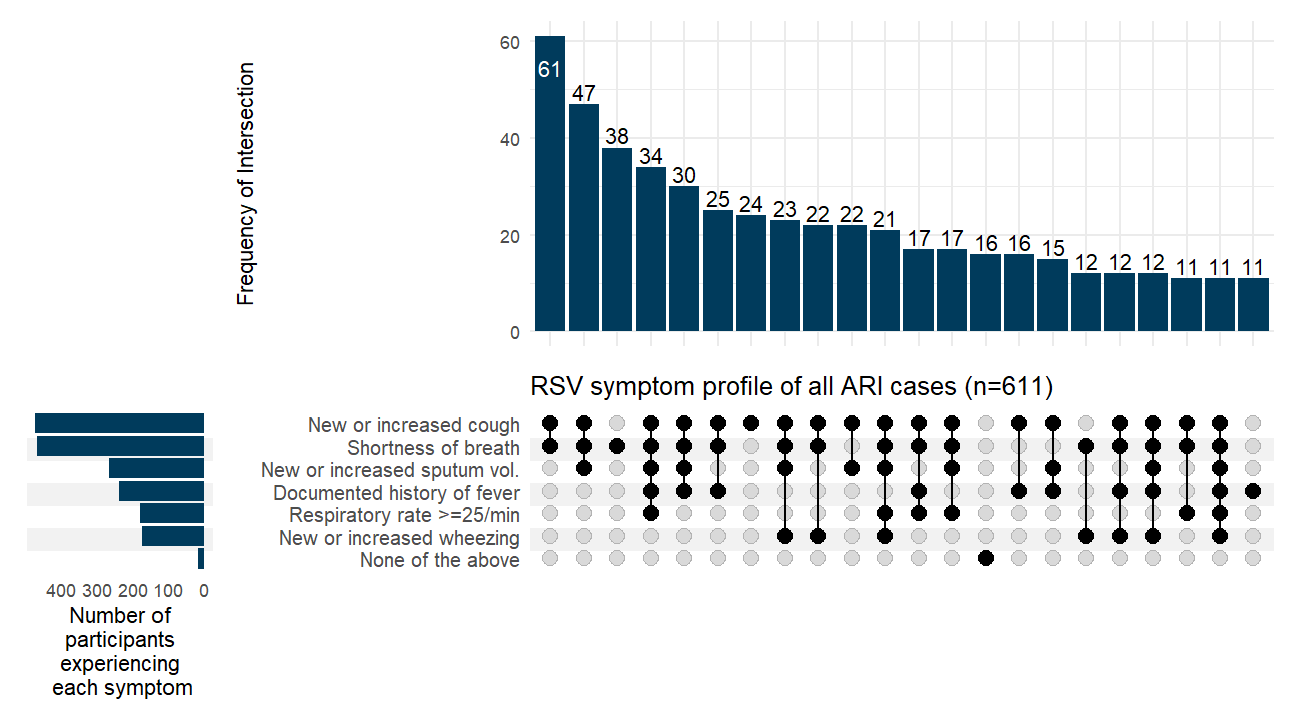

b) **Symptom upset plot for Influenza-associated ARI cases**

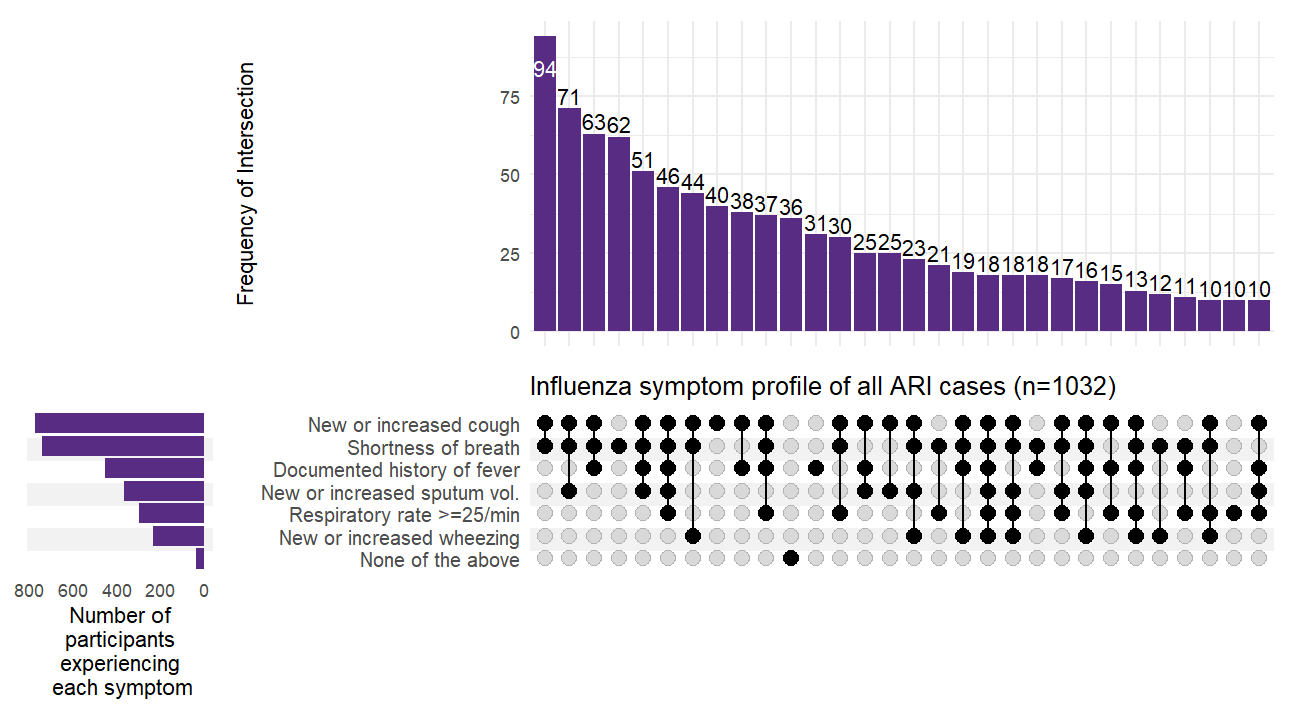

c) **Symptom upset plot for test-negative ARI cases**

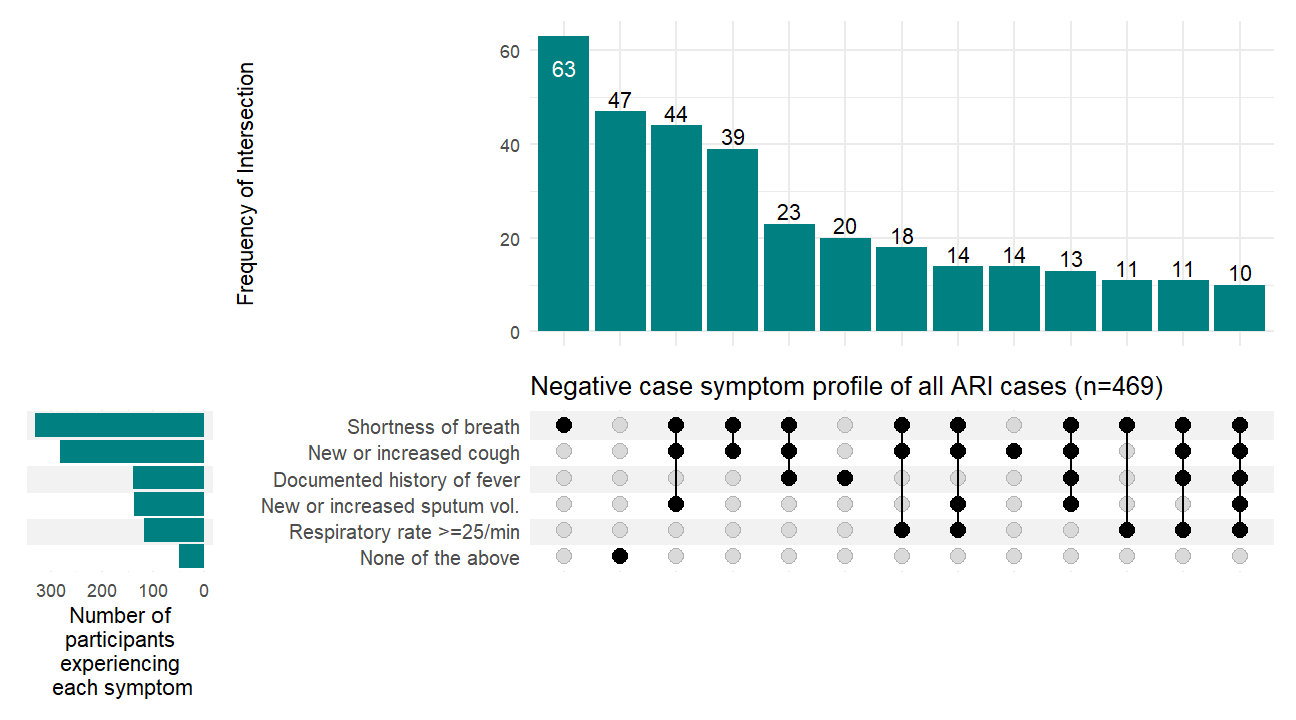

**Supplementary material: Data collection questionnaire**

|  | **Questionnaire** |
| --- | --- |
|  | NHS number, name, date of birth |
|  | **Date of admission** |
| **1** | Date of admission (please enter the date of presentation to hospital for this episode of illness) |
|  | *Calendar box to enter date* |
|  | **Symptoms** |
| **2a** | Did the patient report or present with respiratory symptoms or fever on admission? |
|  | *Yes/No/Unknown* |
| **2b/c** | Date of onset of first respiratory symptoms or fever (or symptoms that led to admission if no respiratory symptoms or fever) |
|  | *Calendar box to enter date* |
| **2b** | Did the patient report or present with any of the following signs and symptoms on admission? (Please tick all that apply) |
|  | *Documented fever (≥37.8 C) or history of fever* |
|  | *Hypothermia (<35.5 C)* |
|  | *New or increased cough* |
|  | *New or increased sputum volume or discolouration* |
|  | *New or increased shortness of breath* |
|  | *New or increased wheezing* |
|  | *Respiratory rate ≥25/min* |
|  | *The patient did not present or report any of the above signs or symptoms on admission* |
|  | *Unknown* |
|  | **Reason for admission** |
| **3a** | Was the patient admitted because of a symptomatic acute respiratory infection? |
|  | *Yes, symptomatic acute respiratory infection primary reason for admission (eg. pneumonia)* |
|  | *Yes, symptomatic acute respiratory infection contributing to admission (eg. RSV infection exacerbating heart failure)* |
|  | *No, admission unrelated to symptomatic acute respiratory infection (eg. cellulitis)* |
| **3b** | The patient was admitted because of… (Please tick all that apply) |
|  | *Pneumonia or pneumonitis* |
|  | *Non-pneumonia lower respiratory infection or acute bronchitis* |
|  | *Exacerbation of chronic lung disease (e.g. COPD)* |
|  | *Exacerbation of chronic heart disease (e.g. heart failure/angina)* |
|  | *Exacerbation of frailty or poor mobility* |
|  | *Symptomatic acute respiratory infection with another reason for admission not specified above (eg. cough syncope) – please specify reason for admission (box provided)* |
|  | **Antiviral use** |
| **4a** | Did the patient receive antivirals (for this illness episode) prior to admission? |
| **4b** | If yes, please specify which antiviral |
|  | *Tick box for: Oseltamivir, Zanamivir, Paxlovid, Sotrovimab, Remdesivir, Other (please specify), Unknown* |
|  | **Severity** |
| **5a** | During this hospital admission, did the patient require any of the following: *(Please tick all that apply):* |
|  | *Oxygen via cannulae or mask* |
|  | *High-flow nasal oxygen (HFNO)* |
|  | *Non-invasive ventilation (NIV) or continuous positive airway pressure (CPAP)* |
|  | *Invasive ventilation or mechanical ventilation* |
|  | *Intensive Care Unit (ICU) admission* |
|  | *Extracorporeal membrane oxygenation (ECMO)* |
|  | *None of the above were required during this hospital admission* |
|  | *Unknown* |
| **5b** | Did the patient die as a result of (primary or contributory cause of death) an acute respiratory infection, or its complications? |
|  | *Yes* |
|  | *No* |
|  | *Unknown* |
| **5c** | If the patient has died, please provide the cause of death as recorded on the death certificate or in the patient notes *(Please enter ‘not known’ if not recorded)*: |
|  | *Cause of death 1a:* |
|  | *Cause of death 1b:* |
|  | **Further information** |
|  | Is there any further relevant information you would like to provide? |
